# A Nomogram to Predict Pneumothorax Requiring Chest Tube Placement following Percutaneous CT-guided Lung Biopsy

**DOI:** 10.1101/2024.02.01.24302030

**Authors:** Masha Bondarenko, Jianxiang Zhang, Ulysis Hugo Baal, Brian Lam, Gunvant Chaudhari, Yoo Jin Lee, Jamie Schroeder, Maya Vella, Brian Haas, Thienkhai Vu, Kimberly Kallianos, Jonathan Liu, Shravan Sridhar, Brett Elicker, Jae Ho Sohn

## Abstract

**Background:** Pneumothorax requiring chest tube after CT-guided transthoracic lung biopsy is one of the common complications, and the required hospital stay after chest tube placement represents an added clinical risk to patients and cost to the healthcare system. Identifying high-risk patients can prompt alternative biopsy modes and/or better preparation for more focused post-procedural care.

**Purpose:** To develop and externally validate a risk nomogram for pneumothorax requiring chest tube placement following CT-guided lung biopsy, leveraging quantitative emphysema algorithm.

**Materials & Methods:** This two-center retrospective study included patients who underwent CT-guided lung biopsy from between 1994 and 2023. Data from one hospital was set aside for validation (n=613). Emphysema severity was quantified and categorized to 3-point scale using a previously published algorithm based on 3×3×3 kernels and Hounsfield thresholding, and a risk calculator was developed using forward variable selection and logistic regression. The model was validated using bootstrapping and Harrell’s C-index.

**Results:** 2,512 patients (mean age, 64.47 years +/-13.38 [standard deviation]; 1250 men) were evaluated, of whom 157 (6.7%) experienced pneumothorax complications requiring chest tube placement. After forward variable selection to reduce the covariates to maximize clinical usability, the risk score was developed using age over 60 (OR 1.80 [1.15-2.93]), non-prone patient position (OR 2.48 [1.63-3.75]), and severe emphysema (OR 1.99 [1.35-2.94]). The nomogram showed mean absolute error of 0.5% in calibration and Harrell’s C-index of 0.664 in discrimination in the internal cohort.

**Conclusion:** The developed nomogram predicts age over 60, non-prone position during biopsy, and severe emphysema to be most predictive of pneumothorax requiring chest tube placement following CT-guided lung biopsy.

## 1. Introduction

CT-guided transthoracic lung biopsy is a widely utilized procedure for diagnostic work up of pulmonary nodules and masses. Although it is considered safe and minimally invasive, the procedure can still be associated with complications, such as pneumothorax, hemothorax, and pulmonary hemorrhage (1,2). Among these complications, pneumothorax is the most common, reportedly occurring in 20-30% of cases (1,3–15). Larger pneumothoraces, occurring between 2-17% of procedures, may require chest tube placement, which is associated with increased healthcare costs and risk of hospital-related infections for patients (1,6–9,12,15–18).

Clinical predictors for complications such as age, gender, patient position, and nodule location have been studied extensively in the literature. The risk associated with emphysema has been reported, although occasionally contested especially in mild to moderate cases (1,3,4,7,8,12,16,17,19–23). Those that have primarily utilized radiologist annotations to classify emphysema presence had significant variability and limited dataset size. Risk models using logistic regression have also been demonstrated in the literature as accurate tools to predict complication, but few have reliably and quantitatively utilized emphysema as a factor.

Some studies have also investigated the use of machine learning models to automatically and reliably classify emphysema extent for the prediction of lung biopsy complication, or directly used chest CT for predictions, but most models were ‘black box’ and/or difficult to translate in a wide variety of clinical settings due to excessive number of covariates (4,16) Part of the debate and variability on identifying predictors of pneumothorax may be attributable many studies having small sample sizes (<1000 patients) and training/validation data originating from a single institution, leading to reduced generalizability of models and results across hospitals.

Therefore, there remains unmet need for a large, multi-institutionally validated algorithm that integrates quantitative algorithm while maintaining simplicity that would allow for immediate clinical usage. Therefore, we aimed to develop and externally validate a risk calculator for pneumothorax complications requiring chest tube placement following CT-guided lung biopsy using a quantitative emphysema algorithm and with emphasis on immediate clinical usability.

## 2. Materials & Methods

### Study Population, Hospitals, and Biopsy Technique Variation

This retrospective, institutional review board approved, informed consent waived, and Health Insurance and Portability Accountability Act compliant study included patients from two hospitals who underwent CT-guided lung biopsy with chest tube placement for pneumothorax management. Cases were from between February 1, 1994, and June 7, 2022, at hospital 1 and between April 16, 2014 and January 30, 2023 at hospital 2. Two hospitals, although affiliated under a single academic institution, had distinct lung biopsy protocols, with mostly distinct primary procedural operators (less than 20% overlapping radiologists across two hospitals). Hospital 1 is a tertiary academic referral center and hospital 2 is a county safety net hospital. The patients at hospital 1 and hospital 2 were mutually exclusive. Operators at hospital 1 utilized the breath hold technique upon needle entry without sedation and with variable use of autologous blood patch at the end of the procedure. Hospital 2 used free breathing technique upon needle entry under moderate sedation followed by saline patch at the end of the procedure.

The ground truth label of chest tube placement was semi-automatically established by matching any patient reports of chest tube placement, if they existed, with reports of CT-guided lung biopsies, followed by manual confirmation.

### Data Preprocessing

Procedural covariates were compiled through report text extraction using both Excel and Python to search for key terms and their variations (e.g. “Right Middle Lobe”, “RML”, “middle lobe”), with missing data from the initial search manually verified and filled in to complete the dataset. Primary reasons for persistent missing data was that CT scans were not accessible, the biopsy was either not a lung biopsy or was a lung biopsy that did not puncture the pleura, and/or the report was missing. Data from hospital 2 was put aside in model development and used for external validation of the logistic regression model.

### Quantitative Emphysema Algorithm

A previously developed and reported quantitative emphysema algorithm was adapted to assess the degree and severity of emphysema in each patient from the biopsy CT scans (24). This algorithm is based on CT image analysis, which uses 3×3×3 kernels and Hounsfield Unit thresholding to determine the extent of emphysema in the lung parenchyma. The algorithm was developed and validated in a separate cohort of patients with known emphysema. Emphysema scores ranged from 0 to 1, and categories were classified into None, Mild/Moderate, and Severe through a weighted logistic regression on various score cutoffs and validation by a trained radiologist. A score less than 0.117^3^ corresponded to no emphysema, a score between 0.117^3^ and 0.215^3^ corresponded to mild to moderate emphysema, and anything above0.215^3^ was considered severe emphysema.

### Variable Selection & Logistic Regression Model

A forward selection method using Akaike information criterion (AIC) with R statistical software (V. 4.2.2) was used to select a combination of variables that would result in the most predictive model. These variables were used to build a logistic regression model in R to predict cases of chest tube placement. The model was developed with data from hospital 1 and validated on data from hospital 2, an external hospital, using bootstrapping and ROC curve analysis.

### Nomogram and Risk Calculator

A nomogram was built from the logistic regression model using RStudio and the ‘rms’ package. A risk calculator was also built and published with R’s Shiny website package using the previously developed logistic regression model.

### Statistical Analyses

Statistical analysis was performed using both R and Python statistical functions. Continuous variables were represented as means ± standard deviations. The association between variables was assessed using Fisher’s exact tests (python scipy.stats *fisher_exact* function) for binary variables, and with a logistic regression (python statsmodel.formula.api *logit* function) for continuous variables. Models were evaluated with bootstrapped calibration curves and Harrell’s C-index. The significance level was set to 0.05.

## 3. Results

### Study Population

A total of 2,512 patients who underwent CT-guided lung biopsy were included in the study, after excluding those with missing imaging and report as well as those with incomplete scans. Of these, 157 (6.7%) developed pneumothorax requiring chest tube as a complication of the procedure, with 1899 patients and 114 complications being from hospital 1 and 613 patients and 43 complications being from hospital 2. The mean age of the hospital 1 cohort was 64.4 ± 14.2 years and 64.6 ± 10.4 years for the hospital 2 validation cohort, and 52% (988/1899) and 59.0% (362/613) were male for the hospital 1 and hospital 2 datasets, respectively. Approximately half of biopsies (hospital 1: 53.23% (1011/1899), hospital 2: 47.47% (291/613)) were performed in the prone position. Further initial covariate distributions are presented in Table 1.

**Table 1:** Summary of Initial Covariates in Both Datasets.

| Risk Factor | Chest Tube in<br>hospital 1<br>Data (n=1899) |  |  | Chest Tube in<br>hospital 2<br>Data (n=613) |  |  |
| --- | --- | --- | --- | --- | --- | --- |
|  | No (n=1785) | Yes (n=114) | P-value | No (n=570) | Yes (n=43) | P-value |
| Age (years) | 64.1 ± 14.3 | 68.2 ± 11.6 |  | 64.5 ± 10.3 | 65.8 ± 11.0 |  |
| < 40 | 124 (6.94%) | 2 (1.75%) | .03 * | 12 (2.10%) | 0 (0%) | 1 |
| Between 40 & 60 | 458 (25.65%) | 22 (19.29%) | .14 | 165 (25.94%) | 11 (25.58%) | .72 |
| > 60 | 1203 (67.39%) | 90 (78.94%) | .009 * | 393 (68.94%) | 32 (74.41%) | .49 |
| Male | 922 (51.65%) | 66 (57.89%) | .20 | 336 (58.94%) | 26 (60.46%) | .87 |
| Non-Prone Position | 812 (45.49%) | 76 (66.66%) | .01 * | 283 (49.64%) | 8 (18.60%) | <.001 * |
| Left Lobe Nodule | 847 (47.45%) | 56 (49.12%) | .77 | 246 (43.15%) | 21 (48.83%) | .52 |
| Emphysema Score | 0.05 ± 0.16 | 0.07 ± 0.17 | .26 | 0.08 ± 0.20 | 0.03 ± 0.05 | .10 |
| Emphysema Extent |  |  |  |  |  |  |
| None | 913 (51.14%) | 42 (36.84%) | .01 * | 318 (55.78%) | 12 (27.90%) | .038 * |
| Mild/Moderate | 355 (19.88%) | 21 (19.42%) | .80 | 82 (14.38%) | 12 (27.90%) | .026 * |
| Severe | 517 (28.96%) | 51 (44.73%) | <.001 * | 170 (29.82%) | 19 (44.18%) | .059 |

\* Significant at an alpha level of 0.05.
Description: Demographics table showing the numeric summary of initial covariates. Continuous variables are shown as mean ± standard deviation. P-values are from Fisher exact tests on binary variables and logistic regression for continuous variables.

### Preprocessing

The flow diagram in Figure 1 demonstrates the dataset creation and preprocessing steps. Not imputable cases with missing nodule location, patient position, and/or gender, totaled to 254 in the training set, and 101 in the validation set, and were ultimately dropped.

**Figure 1:**
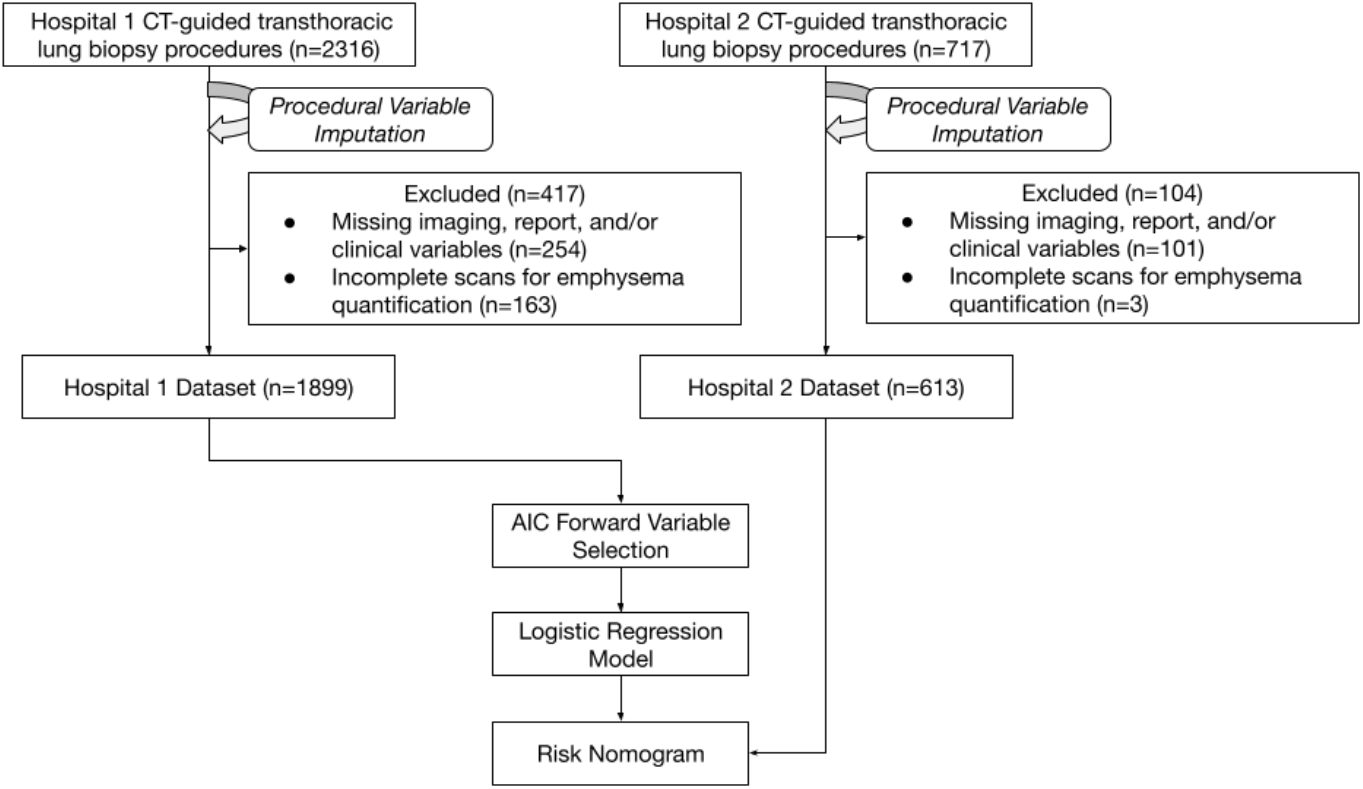
Flow diagram shows patients who underwent CT-guided transthoracic lung biopsy procedures. AIC = Akaike information criterion

### Quantitative Emphysema Algorithm

The emphysema algorithm successfully completed its analysis on all but 163 cases in the training set and 3 in the validation set. These cases were dropped per the exclusion criteria. The percent of patients with none, mild/moderate, and severe emphysema were 50.28% (955/1899), 19.79% (376/1899), and 29.91% (568 /1899) in the training dataset. In the validation set, this corresponded to 53.83% (330/613), 15.33% (94/613), and 30.83% (189/613).

### Variable Selection & Logistic Regression Model

Through multivariable stepwise forward AIC, the following variables were considered in the starting model: male, nodule location left lung, prone position, non-prone position, no emphysema, mild to moderate emphysema, severe emphysema, age less than 40, age between 40 and 60, age over 60, and emphysema score. Age over 60 (OR 1.8 [95% CI: 1.1-2.9], P=.01), non-prone patient position (OR 2.4 [1.6-3.7], P<.001), and severe emphysema (OR 1.9 [1.3-2.9], P<.001) were found to be the best combined predictors of complication. These variables were then fed into the logistic regression model, which achieved a Harrell’s C-index of 0.664 and 0.715 for the training and validation datasets, respectively. A summary of these multivariable findings is presented in Table 2.

**Table 2:** Multivariable Analysis after Model Selection.

| Feature | Odds Ratio | [95% CI] | P-value | Coefficient |
| --- | --- | --- | --- | --- |
| Intercept | 0.01 | [0.01, 0.03] | < .001 * | -3.95 |
| Non-Prone Position | 2.4 | [1.6, 3.7] | < .001 * | 0.91 |
| Severe Emphysema | 1.9 | [1.3, 2.9] | < .001 * | 0.69 |
| Age > 60 | 1.8 | [1.1, 2.9] | .01 * | 0.59 |

\* Significant at p-level of 0.05.
Description: Table presents the results from the final logistic regression multivariable analysis performed on the training data (n=1899). Final variables were selected through Akaike Information Criterion (AIC) forward variable selection from the following factors: male, left lung nodule, age under 40, age between 40 and 60, age over 60, continuous emphysema score, no emphysema, mild/moderate emphysema, severe emphysema, and non-prone position. Emphysema categories were determined by thresholds on values from a quantitative emphysema algorithm. Non-prone position, severe emphysema, and age over 60 provided the strongest model.

There were 34 cases (34/1899; 1.79%) within the hospital 1 dataset and 11 cases (11/613; 1.79%) within the hospital 2 dataset that were false negatives. There were 809 (809/1899; 42.60%) and 297 (297/613; 48.45%) false negatives in the hospital 1 and hospital 2 datasets, respectively. Supplemental Table 3 presents a summary of false negative cases for both datasets.

The model’s calibration was confirmed on the bootstrapped calibration plot, which can be found in Figure 3.

**Figure 2:**
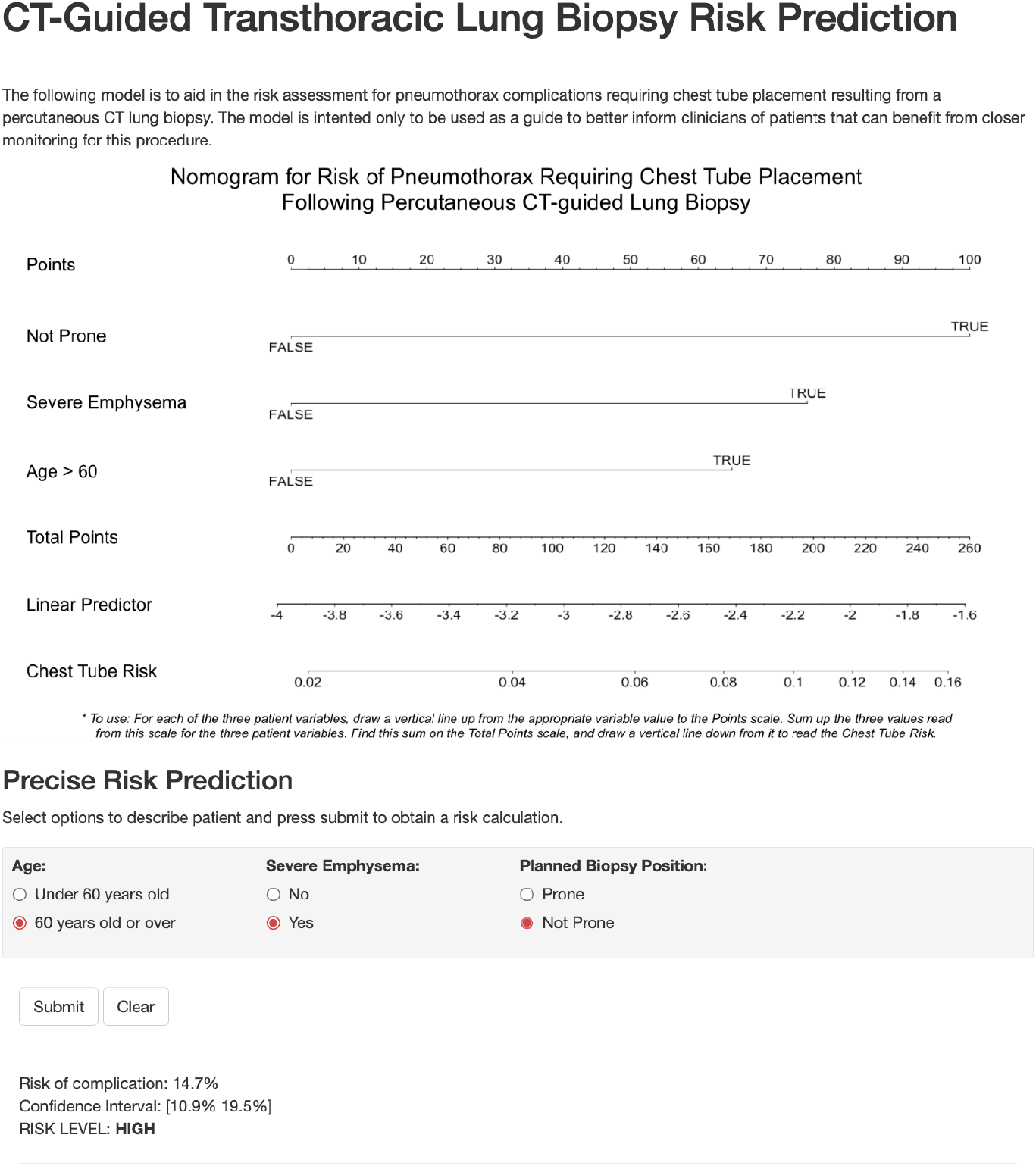
Screenshot from risk calculator website showing nomogram and risk calculator, with an example prediction. Website is publicly available at https://bit.ly/3n9Yu38.

**Figure 3:**
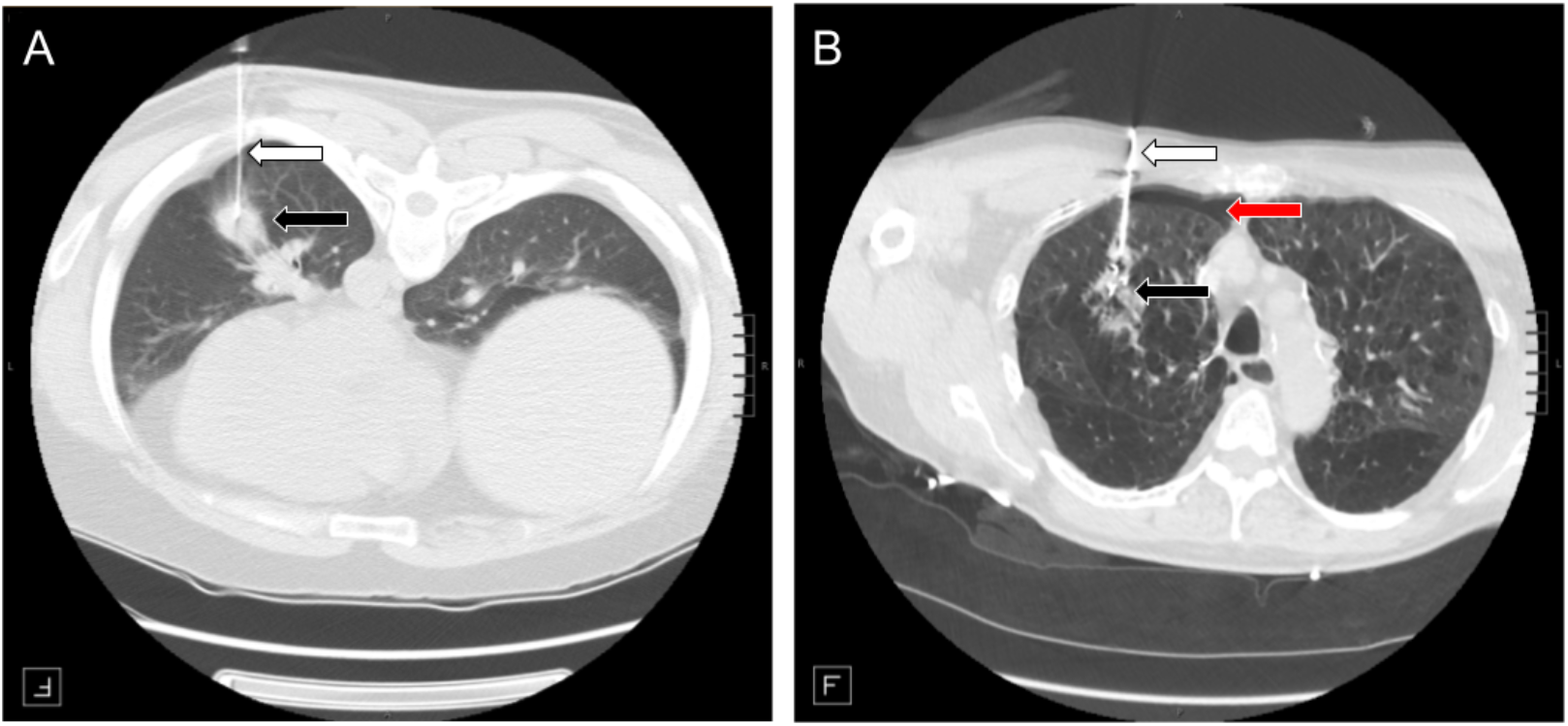
Example cases. CT scans from CT-guided transthoracic lung biopsy in two patients. Biopsy needle (white arrow) and the nodule biopsied (black arrow) are shown. **A)** Patient is a male in their 40s presenting with no emphysema (as determined by quantitative emphysema algorithm), lying in the prone position, with a nodule in the left lower lobe. Our risk nomogram and calculator predicted this patient to have low risk of pneumothorax requiring chest tube (1.86% [1.13% - 3.13%]). The patient post biopsy had a tiny pneumothorax and did not need chest tube placement. **B)** Patient is a male in their 60s with severe emphysema (as determined by quantitative emphysema algorithm), lying in the supine position, and with a nodule in the right middle lobe. Our risk nomogram and calculator predicted this patient to have high risk (14.70% [10.90% - 19.50%]). The patient developed intraprocedural pneumothorax (red arrow) which grew into a large pneumothorax and chest pain necessitating a chest tube placement.

### Nomogram and Risk Calculator

Based on the logistic regression model, a nomogram and a risk calculator (https://bit.ly/3n9Yu38) were developed to predict the risk of pneumothorax with a threshold for positivity at 0.06, which was the average positivity rate for the datasets. Figure 2 displays the nomogram and presents a screenshot of the website. The risk calculator classified patients into low, moderate, and high-risk groups based on their emphysema extent, age, and position during the biopsy. The thresholds for these categories were based on the median (4.562%) and the third quartile (7.953%) of the risks of the hospital 1’s patients as determined by the model. This corresponded to the proportion of patients in the low, moderate, and high-risk groups for hospital 1 data to be 41.4% (788/1899), 23.4% (445/1899), and 35.0% (666/1899), respectively. This proportion was generally maintained in the validation set with the proportion of patients in the low, moderate, and high-risk groups being 33.4% (205/613), 26.9% (165/613), and 39.64% (243/613).

Generally, hospital 2 had patients with higher risk than hospital 1. A distribution of risk predictions for both datasets is graphed in Supplemental Figure 4, and a demonstration of risk groups can be found in Table 3. Figure 3 presents example cases and predictions.

**Table 3:** Risk Groups Stratification.

| Age > 60 | Severe Emphysema | Not Prone | Risk Percent [95% CI] | Risk Group |
| --- | --- | --- | --- | --- |
|  |  |  | 1.89% [1.13% - 3.13%] | LOW |
| ✓ |  |  | 3.36% [2.31% - 4.85%] | LOW |
|  | ✓ |  | 3.70% [2.19% - 6.20%] | LOW |
|  |  | ✓ | 4.56% [2.94% - 7.01%] | LOW |
| ✓ | ✓ |  | 6.50% [4.47% - 9.35%] | MODERATE |
| ✓ |  | ✓ | 7.95% [5.97% - 10.50%] | MODERATE |
|  | ✓ | ✓ | 8.73% [5.52% - 13.50%] | HIGH |
| ✓ | ✓ | ✓ | 14.70% [10.90% -19.50%] | HIGH |

Description: This table presents the predicted risk and corresponding risk group for various combinations of variables in the context of pneumothorax requiring chest tube.

Error analysis of false negative cases by thoracic radiologists demonstrated no identifiable systematic biases.

## 4. Discussion

Our study investigated factors predictive of pneumothorax requiring chest tube placement following CT-guided lung biopsy, leveraging a large dataset (>2000), quantitative emphysema algorithm, and external validation cohort. After pruning the model to simplify for ease of clinical use, we found that non-prone position, severe emphysema, and age over 60 successfully categorized pneumothorax risk and built an online risk nomogram for immediate clinical usage. The risk calculator achieved Harrell’s C-index of 0.664 and 0.715 in the testing and validation cohorts, respectively.

Our risk factor findings are primarily consistent with several previous studies, specifically in regards to non-prone position (22,23,25) and older age (1,3,14,16). Geraghty et al.’s study has also found discretized age, specifically at a boundary of 60 years old, to be statistically significant while age, when represented as a continuous variable, was not. With patient position, other studies have found similar results; non-prone, supine (a non-prone position), and lateral position (a non-prone position) were risk factors from the studies of Nakamura et al. Takeshita et al., and Ruud et al., respectively.

Regarding emphysema, our findings contribute to research that has found presence of emphysema, and more specifically severe emphysema, to be a strong predictor for lung biopsy complications requiring a chest tube. Kazerooni et al. presented an early study that generalized severe obstructive lung disease, as determined by pulmonary function laboratory testing and the American Thoracic Society guidelines, as being statistically significant to chest tube placement. The study by Laurent et al. further claimed severe emphysema, using the same tools and guidelines as the previous study, was significantly associated with chest tube placement. These studies were limited by a moderate sample size and lacked external validation, so our larger sample size and external validation serve to validate these findings.

Our 3×3×3 kernel-based quantitative emphysema algorithm, followed by thresholding to classify into a 3-point category (none, mild-moderate, and severe emphysema) contributed to the reliability and predictive power of the model. Previous studies that leveraged quantitative emphysema was classified using a Hounsfield Unit (HU) thresholding where anything between -950 and -850 HU would be used to calculate percent emphysema in the lungs. This is a simplified version that in our past work appears to overcount the area of emphysema and thus can be prone to error. Theilig et al.’s study found a significant association between these two variables, yet Chami et al. and Lendeckel et al. both did not. These studies had moderately sized samples, all having less than 400 biopsy cases. Our study brings forth once again, using a larger dataset and external validation, findings that quantitative emphysema scores classified into 3-point scale are strongly associated with pneumothorax risk.

Immediate clinical usability was a priority goal in developing this proposed risk nomogram. Multiple previously reported deep learning algorithms as well as traditional machine learning algorithms suffered from well-known black box problem as well as excessive number of variables that resulted in difficulty of clinical integration. We aggressively pruned the model to narrow down the variables to just four easily usable covariates, achieving simplicity and also possibly improving generalizability from model regularization. Even though the quantitative emphysema algorithm was trained using an algorithm, the code is made fully public and most importantly, the thresholding was done and calibrated to radiologist’s classification of emphysema severity. Therefore, the model can take in a qualitative emphysema classification by radiologist as one of its covariates instead of having to exclusively rely on the quantitative algorithm.

By using this tool, clinicians can improve patient management by sending patients to tertiary medical centers or exploring alternative modes of lung biopsy (e.g. bronchoscopic route of biopsy) to mitigate the risk. At minimum, a more informed discussion can happen with the patient to weigh the pros and cons of proceeding with the biopsy at all if potentially higher risk of pneumothorax is predicted.

A few limitations of our results are the study’s retrospective nature, as it could involve conflating decisions made by clinicians to reduce patient risk of complication, as well as the varying decision for when to place a chest tube. The former issue might have resulted in less cases of complication in high-risk patients, resulting in false positives in our risk calculator, while the latter issue might have resulted in differences in complication severity amongst patients that received a chest tube.

In conclusion, we propose an easy-to-use risk nomogram of substantial pneumothorax following lung biopsy integrating older age, emphysema severity, and patient position. Future studies can include additional external validation and calibration at diverse hospital sites and integration of more advanced image processing of the lung parenchyma to improve the prediction.

## Supporting information

supplemental-analyses

## Data Availability

Data generated by the authors or analyzed during the study are available at University of California, San Francisco Information Commons (https://informationcommons.ucsf.edu/).

## Notes

### Competing Interest Statement

The authors have declared no competing interest.

### Funding Statement

This study did not receive any funding.

### Author Declarations

IRB of University of California San Francisco (UCSF) gave ethical approval for this work.

