## supplemental-analyses for "A Nomogram to Predict Pneumothorax Requiring Chest Tube Placement following Percutaneous CT-guided Lung Biopsy"

**Supplementary Figures**

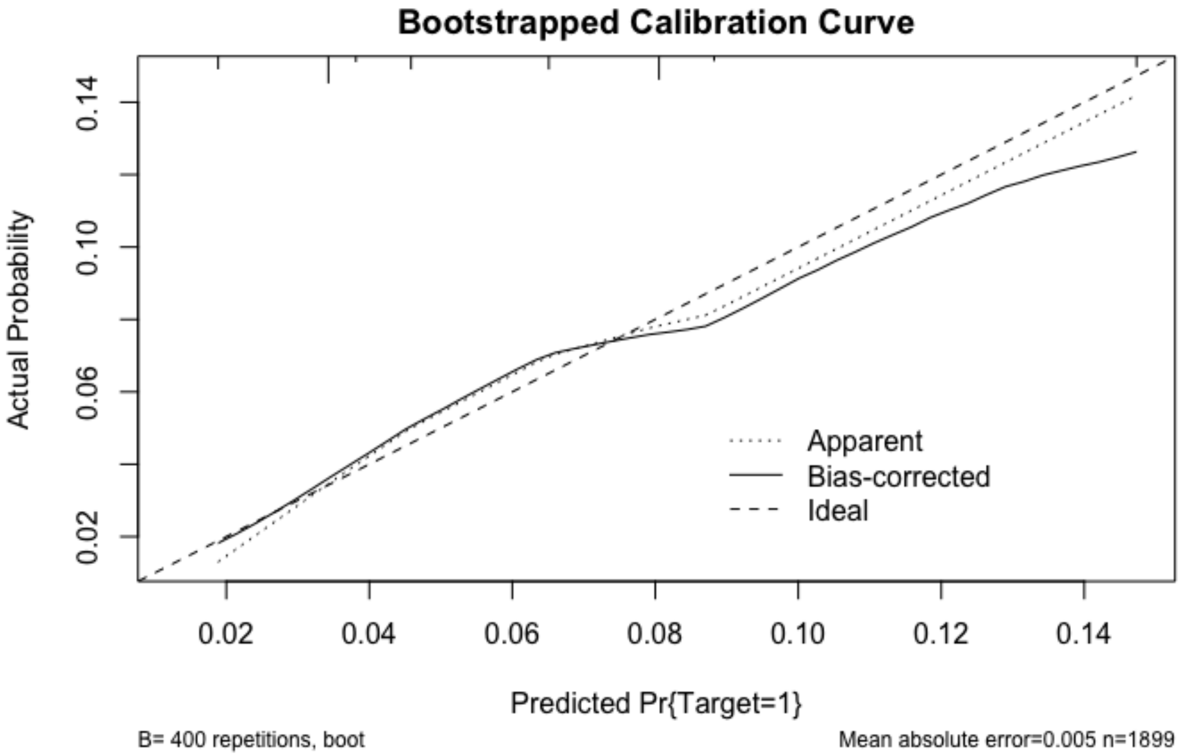


**Supplementary Figure 1:** Figure shows bootstrapped calibration plot for the training dataset, where 'Target' refers to cases of pneumothorax requiring chest tube. 400 repetitions were run to create apparent, bias-corrected, and ideal model lines. A perfect model has apparent and bias-correct lines closer to the ideal line. Based on this calibration plot, our model appears to be well calibrated, with a mean absolute error of 0.005.

**Supplementary Table 1:** Error Analysis on Both Cohorts

Table 1a: Hospital 1 Error Analysis

| Number of Cases | Severe Emphysema | Non-Prone | Age > 60 | Risk Score | Chest Tube Predicted | Chest Tube Needed |
|---|---|---|---|---|---|---|
| 207 | ✓ | | ✓ | 6.49% | Yes | No |
| 12 | | ✓ | | 4.56% | No | Yes |
| 2 | | | | 1.88% | No | Yes |

Table 1b: Hospital 2 Error Analysis

| Number of Cases | Severe Emphysema | Non-Prone | Age > 60 | Risk Score | Chest Tube Predicted | Chest Tube Needed |
|---|---|---|---|---|---|---|
| 85 | ✓ | | ✓ | 6.49% | Yes | No |
| 4 | | ✓ | | 4.56% | No | Yes |
| 3 | | | ✓ | 3.35% | No | Yes |

Description: This table summarizes the false negative and false positive cases for both cohorts. The first row for each table represents the most extreme false positive, the second row shows the most extreme false negative, and the last row shows an additional false negative subset of cases. Hospital 1 had 34 and hospital 2 had 11 false negative cases. There were 809 and 297 false positive cases in Hospital 1 and hospital 2 datasets, respectively.

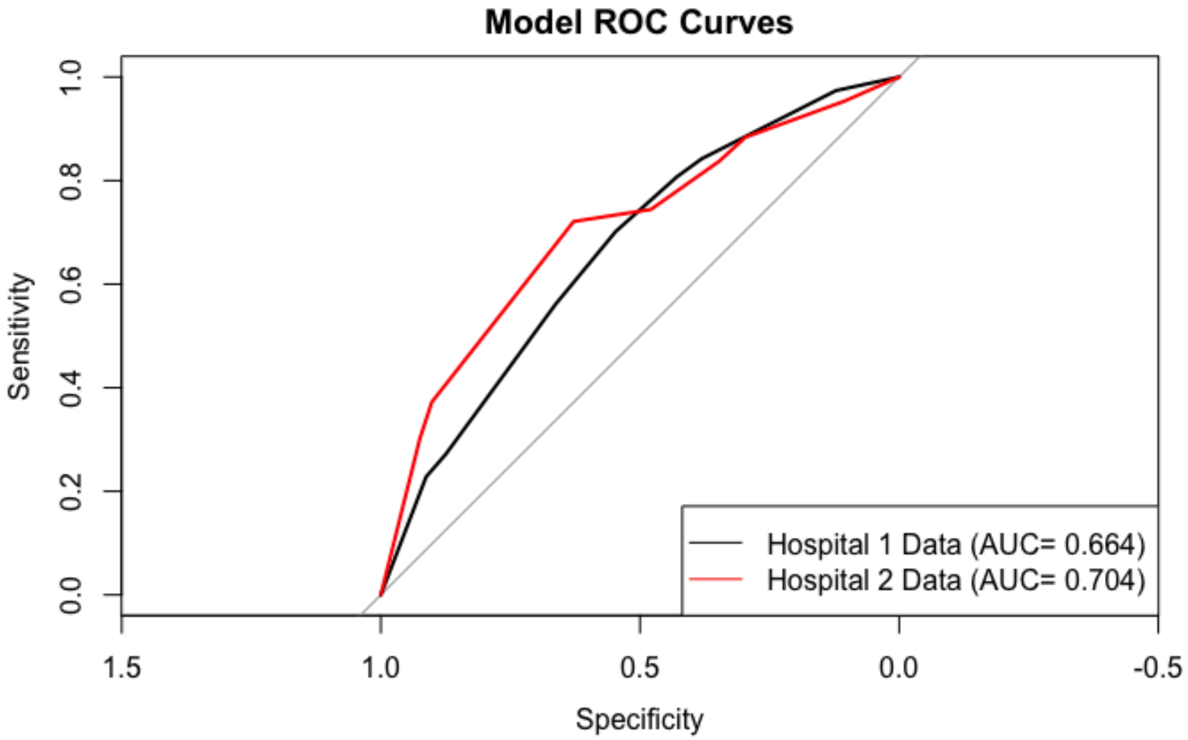


**Supplementary Figure 2:** ROC Analysis shows curves for both training and validation datasets, with the validation curve being higher than the training curve.

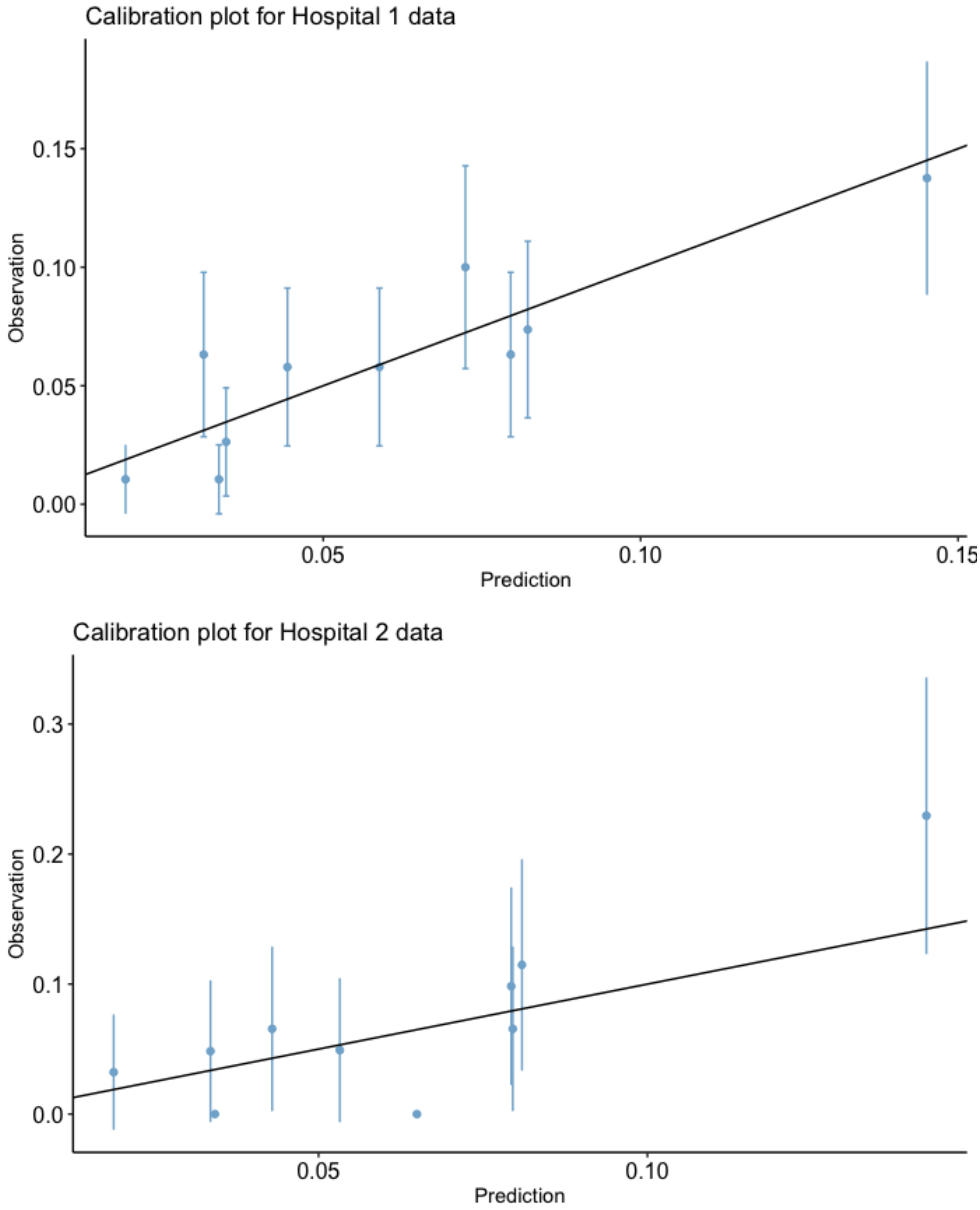


**Supplementary Figure 3:** Figure shows calibration plots for both cohorts.

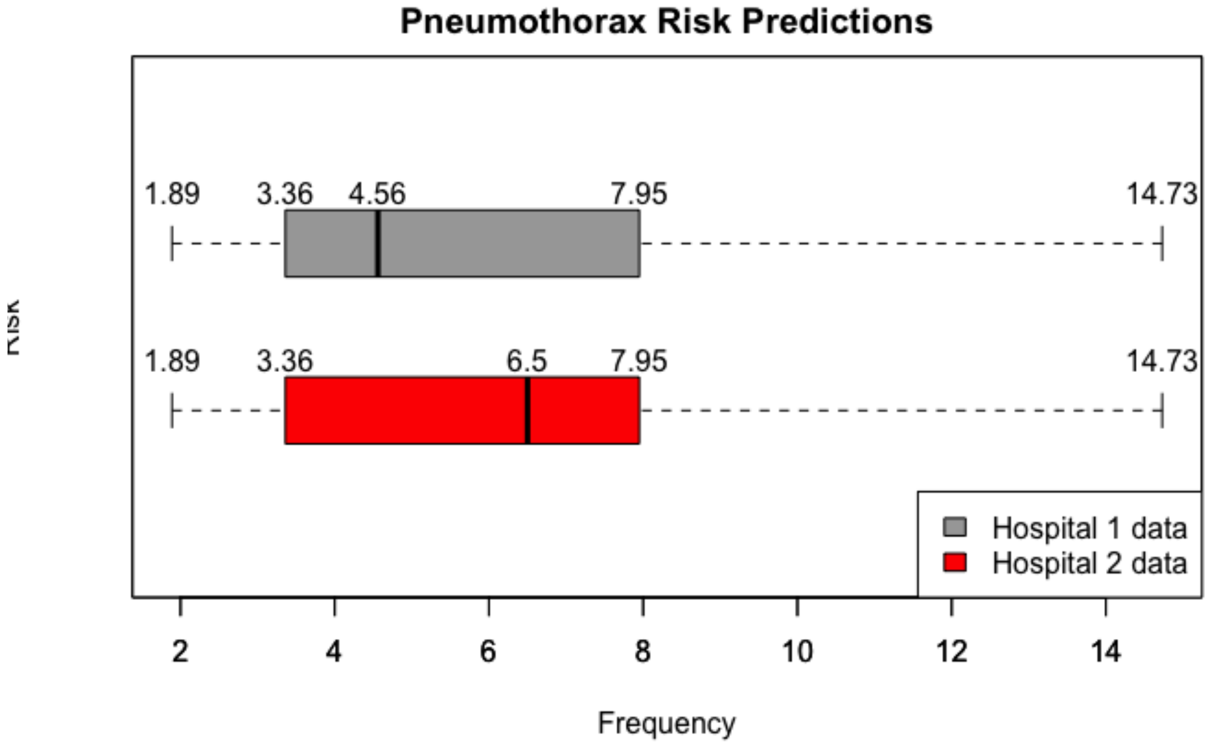


**Supplementary Figure 4:** Figure shows distribution of risk scores for both training and validation datasets. Figure shows both datasets have the same range and quartiles of predictions, but with different medians. The validation dataset had the higher median.
